# Practices, perceptions, and ethical concerns of antimicrobial use among poultry farmers in Kiambu County, Kenya. One-Health context

**DOI:** 10.1101/2024.10.15.24315541

**Authors:** Ann Munene, Hamilton Majiwa, Elizabeth Bukusi

## Abstract

Globally, the poultry sector is one of the primary animal protein sources for human consumption. The poultry sector enhances both food and economic security in Kenya. This has led to adaptations in the poultry sector to respond to the growing demand for animal protein. Practices such as antimicrobial use for disease management, growth promotion, and product improvement within the poultry industry have led to public health, environmental, and ethical concerns. A predominant poultry-producing region in Kenya, Kiambu County was chosen for this study. In-depth interviews (n=21) were conducted until saturation among both small- and large-scale poultry farmers of layers, broiler, and indigenous chicken breeds. The farmers revealed overuse and misuse of antimicrobials, use of antimicrobials for growth promotion and product improvement, and non-adherence to withdrawal periods. They also use poultry waste in other animal feeds and improperly dispose of antimicrobial dilutions. These practices are all likely to contribute to the development and spread of antimicrobial resistance (AMR), which is a global threat to human, animal, and environmental health. The lack of awareness of the potential harm caused by the practices and disregard of instructions, demonstrates the need for increased awareness among poultry farmers on rational use of antimicrobials in poultry farming. Poultry farmers’ ignorance and lack of adherence to regulations of antimicrobial use in poultry farming raises ethical concerns about the threat to human, animal, and environmental health. We conclude that a multifaceted One Health approach is required to evaluate the different ethical, social, and biological factors that contribute to the development and spread of antibiotic resistance arising from poultry production to safeguard public and environmental health.

## Introduction

Human, animal, and environmental health are intertwined and One Health research estimates that 60% of human pathogens are zoonotic. Of more than 30 new human pathogens detected in the last 3 decades, 75% are of animal origin (1). Globally, animals that are meant for food offer livelihood, food and nutrition security, and economic value (2). In Kenya, the poultry sector is one of the pillars of food and economic security (3). Practices within the industry have contributed to the emergence of public, environmental, and ethical concerns (4). Worldwide, antimicrobial use (AMU) is highest in poultry farming compared to other animal production systems (3, 5). This contributes to antimicrobial-resistant bacteria found in poultry, humans, and the environment (6). Antimicrobial resistance (AMR) refers to a state where a microbe is no longer susceptible to an antimicrobial that it was previously susceptible to (7). AMR remains a growing global risk to human, animal, and environmental health (8). Recent studies reveal estimates of annual global deaths attributable to AMR by 2050 will reach 10 million (9), with Kenya being listed as a new AMR hotspot (10). A high risk of AMR is associated with small scale, unregulated poultry farming operations in low-income settings (11). In Kenya and globally, the poultry chain is a current concern regarding the spread of antimicrobial resistant infections. Antimicrobial misuse in the poultry sector has contributed to therapeutic failure of medicines intended for both human and animal health, posing a health risk to humans, animals and environment (3, 11). Unethical practices continue to promote antimicrobial resistance in One Health.

Ethics, derived from the Latin word "ethos" denoting character, involves a collection of behavior or actions constrained by a set code principles or moral standards that advance the interests of humans, animals and the environment (12). A fundamental idea in medicine is the ethical principle of nonmaleficence, which conditions do-no-harm (13). Inappropriate antimicrobial use may have unintended consequences, such as the development and spread of resistant bacteria and exposure of antimicrobial residue to animals, humans, and the environment. This calls for a holistic approach to AMR to preserve the efficacy of antimicrobials for future generations (14). In humans, unethical practices such as antimicrobial self-prescription without doctors’ advice, over-the-counter dispensation of antimicrobials by pharmacists without a doctor’s prescription, or laboratory diagnostic results, inequitable access to antimicrobials, and a lack of adherence to treatment regimes, are among the practices that contribute to AMR (15). Of growing concern is the justification for use of antimicrobials in animal production defended by the claims of improving animal health and welfare for the availability of food for a growing population (16). This has led to unethical practices, such as incorporating antimicrobials for growth promotion and as feed efficiency enhancers, exposing humans and the environment to potentially antimicrobial-resistant pathogens and antimicrobial residues. Although the use of antimicrobials in animal and agriculture production contributes to economic gain, the risks associated with overuse and misuse of antimicrobials outweigh the benefits (17). The reservoirs and resistant genes in the environment and poultry products, such as egg yolk, arising from antimicrobial use in poultry pose a public health hazard to consumers (18, 19). Antimicrobials used in human health including last resort antimicrobials such as colistin have been applied to boost animal output (20, 21). Non-adherence to antimicrobial dosage and recommended withdrawal periods exposes poultry consumers and the environment to antimicrobial residues (22). Many farmers are hesitant to adhere to withdrawal periods citing economic losses arising from discarding poultry products when the poultry are on treatment, therefore raising some ethical concerns about intentional exposure of potential harm to consumers of poultry products (18).

The environment is also more vulnerable to the development of resistant microbial genes and antimicrobial residues when poultry waste is improperly disposed off, including the application of litter as manure (23). AMR is both a social and biological challenge and requires a socio-economic solution. There is a paucity of data regarding ethical concerns and awareness of antimicrobial use among poultry farmers in Kenya. Knowledge, attitudes, and practices of antimicrobial use and antimicrobial resistance are critical aspects for consideration to promote the ethical use of antimicrobials and address the global threat of antimicrobial resistance (24). There is a need to understand practices and attitudes to advocate for antimicrobial stewardship under a One Health approach, including policy formulation and implementation to reduce harm arising from appropriate and inappropriate use of antimicrobials (17).

## Methodology

### Study site

The study was conducted in Kabete and Kikuyu sub-counties of Kiambu County, central Kenya. After Nairobi County, Kiambu County has the second-highest population of approximately 2,417,735 people (25). Kiambu is known for its abundant rainfall and rich soils with a lot of promising smallholder farms that can produce poultry and dairy products, fresh fruits, and green vegetables to feed the county as well as neighboring counties. The greatest population of commercialized chicken and the highest use of antimicrobials in poultry production has been documented in Kiambu County (3).

### Study Design

This research employed a cross-sectional exploratory qualitative approach due to its flexibility and ability to best address the research question. In-depth interviews were used to obtain qualitative data, between 25th March and 5th April 2024. Research on knowledge, perceptions, and practices were broadly investigated to identify behavioral patterns and knowledge among poultry farmers to effectively document and explore One Health interventions and ethical use of antimicrobials in poultry farming.

### Study population and unit analysis

The study population comprised of poultry farmers of indigenous, layer, and broiler chickens. Farmers with more than 500 chickens were considered large scale, while those with more than 50 but less than 500 were denoted small scale (26). The total sample size was twenty-one (n=21), among them seven males and fourteen females. The unit of analysis was the individual poultry farmer.

## Sampling procedure

### Field entry

A connection with the poultry farmer’s community was established through the respective sub-county veterinary officers who serve as leaders of veterinary services. This was further enhanced through a linkage between the poultry livestock production officers who helped in mapping the target farmers. Livestock production officers work under the veterinary officers as a link between the farmers and the government services and support the coordination and dissemination of livestock production information to farmers. A meeting was held between the principal investigator (AM), the sub-county veterinary officer, and the livestock production officers to explain the study before meeting the study participants and ensuring they understood that it was an entirely academic study with no ulterior financial gain. The livestock production officers helped to generate and map a poultry farmers list for each sub-county sampled. A farmer had to be over the age of eighteen and have raised chickens for more than two years. This inclusive criterion was to allow for at least one complete cycle of production of layer chicken up to the disposing stage. Farmers who met the inclusion criterion were numbered and mapped on the list of the poultry farmers supplied by the sub-county offices. Study participants were randomly selected from the list using odd numbers. Knowledgeable and experienced participants in poultry keeping were interviewed on the study topic to maximize the likelihood of gathering and providing valuable and relevant data. The principal investigator walked each research participant through the informed consent process and giving them a comprehensive explanation of its contents to ensure that they understood that participation in the study was voluntary. After explaining the purpose of the study to them and obtaining their permission through a signed informed consent form, the participants were interviewed in a quiet area on the farm.

### In-depth interviews

To allow the informants to carry on with their business without significant interference, the in-depth interviews were conducted at their farms using a voice recorder for audio recording and a structured guide that allowed for probing. This method was crucial in digging out data on the respondents’ knowledge, perceptions, and practices exploring ethical concerns surrounding antimicrobial use in poultry farming. The method’s semi-structured design played a crucial role in identifying ethical concerns related to certain practices. The approach produced unprompted answers that were important to the investigation. Additional in-depth interviews with respondents were conducted up to the point of saturation when their responses provided no more new themes, concepts, opinions, or patterns (27).

### Data processing analysis

Thematic analysis, a method for identifying, analyzing, organizing, and describing themes within a data set, was employed (28). Once the qualitative data was obtained the audio data was transcribed, with a translation into English in cases where the interview was conducted in Swahili, the national language. Data category and coding were done after verbatim transcription. Transcription involves converting recorded audio to a written format. The transcripts were uploaded into Nvivo and a codebook was developed for deductive analysis, in which a predetermined set of codes was applied to the data (29). The transcripts were read in detail and coded. Subsequently, the data was subjected to a thematic analysis, which involved creating themes consistent with the goals of the study by picking out emergent patterns in the informants’ responses. Deductive coding focused on relevant themes which included the use of antimicrobials, adherence to treatment, withdrawal period, use of poultry waste, and access to veterinary and extension services. The study examined respondents’ perceptions, practices, and ethical concerns regarding the use of antimicrobials in poultry keeping. Some direct narratives and quotations have been presented in the findings of the study.

### Ethical consideration

The study was conducted in compliance with good research standards and laws by taking the required ethical considerations and actions. Ethical approval was sought from the Biosafety, Animal Use and Ethics Committee, Faculty of Veterinary Medicine, University of Nairobi (Ref: FVM BAUEC/2024/538). A research permit was granted by the National Commission for Science, Technology, and Innovation (NACOSTI) (Ref no: 746760). Each participant gave written informed consent for their participation in the study and for the audio recording of their interview.

## Results

A total of 21 study participants, (seven males and fourteen females) were interviewed. The socio-demographics of the research participants are presented in Table 1.

**Table 1:** Demographic profile of the respondents.

| CHARACTERISTIC | FREQUENCY<br>n=21 |
| --- | --- |
| Gender |  |
| Male | 7 |
| Female | 14 |
| Age |  |
| 25-34 | 1 |
| 35-44 | 5 |
| 45-54 | 4 |
| 55-64 | 5 |
| 65-74 | 4 |
| Over 75 | 1 |
| No age recorded | 1 |
| Education level (Kenyan education system) |  |
| Secondary | 11 |
| College/Tertiary | 10 |
| Poultry type |  |
| Layers | 12 |
| Layers, Broilers | 2 |
| Layers, Broilers, Indigenous | 3 |
| Indigenous | 3 |
| Broilers | 1 |
| Type of farm |  |
| Small scale 50<500 | 10 |
| Large scale > 500 | 11 |

Most of the poultry farmers were women and between the ages of 35 and 60, with the youngest being under thirty and the oldest being over seventy-five years of age. They kept three types of individual breeds of chicken: layers, broilers, indigenous, and/or a mix of the types. The layer breed type was predominant among both the small- and large-scale farmers in the region. Farm sizes sampled at the large and small scale were distributed equally. Education attainment was generally high where all the participants had post-primary education with each half having exposure to either secondary or tertiary education.

### Emerging themes

To find out more about the informants’ understanding of and actual antimicrobial use, questions posed to them investigated their knowledge of the effects of antimicrobial use on animals, humans, and the environment. The study identified six themes of ethical concern.

## 1. Use of antimicrobials in the absence of disease

Farmers admitted that they use antimicrobials along with other treatment options, like vaccinations and herbal concoctions, during poultry keeping. Some farmers admitted that it was difficult to avoid using antimicrobials in poultry production. Quotes are italicized showing varied responses from the farmers.

> **Large scale Farmer**: *“Yes, I have always used antimicrobials. Some drugs are mandatory, whereby you have to administer them to the chicken whether there’s an ailment or not. Some I administer only when my birds are ailing while others are just for prevention for future illness”*.

However, poultry farmers who kept indigenous chicken opted for other options and avoided using antimicrobials on their poultry as quoted below.

> **Small-scale farmer of layers and indigenous chicken:** *“I don’t like giving them antimicrobials because they develop resistance. I’m trying to make them develop natural immunity. That’s why I prefer using Aloe Vera and pepper but if need be, I may give them medication once in a while.”*

> **Indigenous chicken Farmer**: *“I prefer natural things, so I don’t give them any conventional medicine”*.

> **2^nd^ Indigenous Chicken Farmer:** *"I dislike administering medication to my chickens. For instance, you could discover that chickens are low in calcium. I then learned that you simply take the egg shells, shell and dry them, and then combine them with the poultry feed. That already includes a calcium supplement. Searching for commercial calcium supplements is not necessary. I prefer to pursue the natural route rather than the artificial one, therefore I add natural ingredients to my chicken feed”.*

## 2. Adherence to treatment regime

Some farmers were aware that irrational use of antimicrobials could lead to antimicrobial resistance while others were not. The majority obtained a prescription from agrovets, the shops that deal with veterinary supplies among other products for the farmers. The study identified variations in dosage calculations and how dilutions were constituted among different poultry farmers. Most farmers stated it was important to adhere to prescriptions and dosage instructions as revealed in the excerpts below:

> **Small -scale farmer:** *“Yes, it is important to follow drug prescriptions. In my case, I estimate the right dose in my water tank for the entire treatment period and my chicken drink the water until the prescribed days are over. I don’t think the manufacturers of these drugs were foolish. If you don’t follow the guidelines as instructed the chickens might form a resistance to the drugs”.*

> **Large-scale Farmer:** *“It’s just like in human beings. When the doctor prescribes a dosage, you must finish it or you could under dose and bring complications on yourself.”*

> **Large-scale Farmer:** *“With such an investment you do not take any risks. Even after recovery, you still have to ensure the dosage is administered completely. I replace the dose every day until they finish the number of day’s treatment required”*

On the other hand, some farmers expressed a different opinion on dosage adherence and stated that they were well informed and it was not necessary to follow the prescription guidelines from the agrovets officials as quoted below.

> **Small scale farmer:** *“If you are informed as I am, you don’t have to do exactly as they say”.*

## 3. Adherence to withdrawal period

Some ethical dilemmas and concerns surrounding antimicrobial use were identified. Some farmers stated that they were informed about withdrawal periods but they did not observe them due to possible financial losses that arise from discarding poultry products when poultry is on treatment. Other farmers argued that it is possible to purchase antimicrobials that do not require a withdrawal period. Several farmers who engaged in breeding layer chicken stated that even if antimicrobials affect the chickens’ meat they do not affect the eggs because eggs are an external product. Various quotes are expressed;

> **Small-scale farmer of the layer breed:** *“You cannot consume the broilers meat for seven days because the drug is still in their veins. The eggs do not have any effect”.*

> **Large-scale farmer:** *“I believe it is essential to observe the withdrawal period due to drug effect. However, it is unrealistic for a farmer to dispose of one’s product just because of the withdrawal period. Most farmers tend to purchase drugs that do not have a withdrawal period.”*

> **Large-scale farmer**: *“Sometimes it is stated that we should observe a withdrawal period before consuming the chicken’s products. This is very hard to follow especially when you are in agribusiness.”*

## 4. Use of poultry waste in crop farming, animal feed, and waste disposal

Farmers articulated that the poultry waste and litter were not harmful and could be used for feeding other livestock and for enhancing crop farming. Different farmers responded as follows:

> **Large-scale farmer:** *“I use the waste for my farm to feed the cows. I sieve it and mix with other feeds like dairy meal and napier grass or other fodder. The mixed feed has a lot of vitamins and is very nutritious for the cow because of the chicken waste.”*

> **Small-scale animal farmer**: *“The poultry waste is reduced because I take the waste to the farm later to grow my crops. Therefore, nothing from my poultry farm is wasted including the waste itself.”*

Some farmers argued that they have been feeding the poultry feed waste to the cow for better production of milk and there was no harm in doing so.

> **Large-scale male farmer:** *“I have been doing this since 1990s and there is no problem. I mix the poultry waste with the cows’ dairy meal and feed them once a day in the evening and twice if they are being milked. This enhances more production of milk to distribute for sale every morning.”*

Most farmers did not have a designated environment for disposing left-over antimicrobials, or waste water containing antimicrobials, after treatment. They stated there was there was no environmental impact from antimicrobial disposal. Some quotes of varied responses include:

> **Large-scale farmer:** *“To be honest, I normally just pour the left-over dilutions outside my compound then I prepare fresh medication. I have never thought it could harm the environment pardon my ignorance.”*

> **Small-scale farmer:** *I pour any remaining medication in the pit latrine*.

## 5. Access to veterinary and extension services

Agricultural extension officers and livestock production officers are government officials who provide farmers with essential advice and knowledge on improving crop and animal productivity. Through community engagement, they also help in the management and control of diseases in crop and animal production among other services. Nearly 90% of the respondents cited the absence of government extension services as being the primary challenge in chicken production. The farmers reported that they hardly ever get government veterinary personnel visits to their poultry farms, instead they rely solely on private veterinary services sourced at personal cost. Some farmers reported that they transported sick or dead birds to the agrovets shops for postmortem analysis and/or guidance on how to treat poultry illnesses. Many farmers stated that training was necessary for managing diseases and the appropriate use of antimicrobials in poultry production. The excerpts below express some views of the farmers:

> **Large-scale farmer:** *“The veterinary doctors only come to the farm when you call them, but you have to pay. They are private practitioners, people who study veterinary medicine and decide to practice privately. In the past, we had government officials from Kabete, but they no longer visit our farms.”*

> **Indigenous chicken farmer:** *“Extension officers employed by the government should put in more effort in distributing poultry keeping information.”*

> **Small scale farmer:** *“I have not been in any training, but I think it’s very important to be trained because that’s how I would gain more knowledge. Maybe they can train me about the important drugs that I have not been using but are necessary. I really need to be trained.”*

> **Large-scale farmer**: *“Companies have been training us about different topics and we like these trainings. However, the government does not. That is why we attend these workshops. It is important that we are trained about drugs and feeds. Feeds are a very big problem because we have very many companies each saying they sell the best.”*

## 6. Knowledge and perception of AMR and AMR residues

Leading questions that prompted in-depth answers from the informants were used to assess knowledge and awareness of antimicrobial resistance. The questions investigated the informants’ actions and awareness of the effect of antimicrobial use on animals, humans and the environment. The majority of the farmers did not realize that their practices and actions could contribute to AMR. Many farmers had the perception that antimicrobials become inactive with time and therefore cannot cause any harm to humans, animals, or the environment after use. Some of the farmers who raised layer breeds claimed because the layers are retained for a longer period, the antimicrobials lose effect over time. In contrast, they further claimed that broiler breeds, which have a shorter farm-to-market timespan, may cause harm to humans when consumed. This is evidenced in the quotes below:

> **Small-scale farmer:** *“The drugs we use are not harmful because the doctors only give us the medication to help us and not to bring harm.”*

> **Large-scale farmer:** “*Poultry drugs cannot harm the environment. For instance, if I pour any leftover medication in the environment, it will eventually decompose.”*

However, few farmers were aware that irrational use of antimicrobials could lead to antimicrobial resistance as quoted below:

> **Indigenous chicken farmer:** *“Antimicrobial resistance is a situation whereby the drugs can no longer cure the chickens. Mostly, caused when you don’t finish a dose.”*

## Discussion

This study evaluated the perceptions and practices that raise ethical concerns about antimicrobial use in poultry farming. The key findings of the study reveal that most poultry farmers use antimicrobials for poultry production and do not follow the recommended withdrawal period during the antimicrobial treatment regime. Approximately, 81% of the farmers interviewed obtained prescriptions from the agrovets shops, however, there are variations in the dosage administration. Poultry farmers in general, were unaware that antimicrobial residue remains present in poultry products. The farmers lacked access to government extension services and training in poultry production.

### Irrational use of antimicrobials

Antimicrobial abuse and overuse in the veterinary, agricultural, and healthcare sectors contribute to the global acceleration of AMR (30). There was evidence of inappropriate use of antimicrobials by both small- and large-scale poultry farmers, especially in the production of layer and broiler breeds for commercial purposes. As evidenced by some poultry farmers, administering antimicrobials without a clear indication of a bacterial infection, and over-the-counter access, are prominent examples of antimicrobial misuse leading to AMR (31). Some of the poultry farmers indicated the use of antimicrobials to improve production, and for prophylaxis as a caution for future illness. Research has shown antimicrobial use in production of food animals ought to be limited to the treatment of diseases that have been validated by a professional veterinarian or, in certain cases, to contain an epidemic of disease, to protect the effectiveness of these life-saving drugs in treatment (32). As stated elsewhere, (33) while boosting animal output, antimicrobial growth promoters should be replaced with safer alternatives that are less hazardous to the animals and the environment, such as probiotics and prebiotics. There were variations in dose administration to the poultry indicating inappropriate use of the antimicrobials. It is critical to follow and correctly apply antimicrobial dose recommendations to prevent treatment failures and the emergence of AMR (34). However, small-scale farmers of indigenous chicken reported the use of alternative treatments ranging from natural remedies to herbal medicines, indicating these types of farmers are likely not to use antimicrobials to rear indigenous chicken. This is consistent with the findings that (35) herbal treatments are used more by domestic farmers while antimicrobial use is more rampant in commercial poultry production.

### Non-adherence to withdrawal period

Human populations may be exposed to veterinary antimicrobial residues through the consumption of animal products even if they are not intended for use on people, jeopardizing human health (36). The study revealed that most farmers were aware of the withdrawal period of chicken products during antimicrobial treatment, but did not adhere to it to avoid financial losses (37). Intentional non-adherence to the withdrawal period of poultry products by poultry farmers raises ethical concerns due to the potential harm it may cause to humans, animals, and the environment (3, 36). However, the farmers raised concerns about how they would be compensated for the loss of the products while observing the withdrawal period, requiring a way to mitigate income risk in agribusiness.

### Knowledge of AMR and AMR residues in poultry products

Knowledge of antimicrobial use is essential to reduce antimicrobial resistance in humans, animals, and the environment. Given the disparities in age and gender among poultry farmers, educational initiatives must be designed to influence all age groups and genders regarding the ethical and logical application of antimicrobials in chicken production (38). Regarding age and gender, it was shown that poultry farming was practiced by males and females cutting across all ages, from the youth to the elderly. Regarding education, most of the participants were able to identify antimicrobial agents and the importance of following prescriptions during treatment. This could be explained by the fact that all the research participants had attained secondary school-level of education, implying that education is essential for raising actors’ understanding and awareness of the appropriate use of antimicrobials (39, 40). However, this study revealed limited knowledge of AMR and AMR residues by most farmers. Their responses indicated that they perceived antimicrobials as useful for treatment and are not harmful to animals, humans, and the environment. Antimicrobial residues and antimicrobial-resistant genes are mainly released into the environment and spread to humans and animals through manure from food-producing animals (41). Moreover, antimicrobial use in animals raised for food may lead to the presence of residues in edible animal products (41). All the poultry farmers interviewed indicated that they use poultry manure in the farms to improve crop farming, and some recycled the poultry manure and feed waste as feed for other animals such as cows and pigs. They claimed that antimicrobials become ineffective with time hence disposing of antimicrobials in the environment has no harmful effect. They claimed that a longer stay on farms of birds, like the layer breeds, before disposal, resulted in a gradually waning antimicrobial effect. Additionally, they associated the presence of antimicrobials in the blood, veins, and body tissues of the birds, but not in the eggs, which are an external product. This perception indicates unawareness of antimicrobial residues in the environment by the poultry farmers as reported elsewhere (42, 43). As documented in other countries, the lack of adequate knowledge and awareness of antimicrobial resistance propels malpractices encompassing ethical use of antimicrobials among poultry farmers, jeopardizing One Health (44, 45, 46, 31). This study clarifies the intricate interactions among pragmatic factors that influence poultry farmers’ actions in relation to antimicrobial use. It highlights the importance of a deeper understanding of these practices to design effective interventions that will promote ethical antimicrobial use in One Health among the community of poultry farmers to curb the threat of antimicrobial resistance.

### Lack of government extension services and training for farmers

In the study, the farmers reported the lack of government veterinary farm visits, and agricultural extension services, including training on best farming practices, such as rearing one-day-old chicks to maturity, disease diagnosis, and management. This agrees with the finding (47) that there is a lack of and a need for veterinary officers and agricultural extension services, including the training and awareness of overuse and misuse of antimicrobials among poultry farmers in the Kiambu region. It implies that most farmers may not know when to use and when not to use antimicrobials in poultry production. Through information sharing, veterinarians and agricultural extension officers can help resolve many of the moral quandaries surrounding the production of animals for food (48). As stated elsewhere, the farmers are compelled to seek veterinary services from agrovets stores and private veterinary practioners. Consequently, this raises ethical questions about the conflict of interest that may arise from pharmaceutical company’s potential desire to market antimicrobials to farmers in absence of bacterial infections even when alternative treatment and disease management options exist (31). As reported elsewhere, antimicrobial resistance cannot be fully addressed by reduced antimicrobial use alone, hence the need for training on ethical use of antimicrobials to bolster the battle against antimicrobial resistance (49).

### Conclusion and recommendation

This study reveals the overall practices that raise ethical concerns about antimicrobial use by poultry farmers in Kiambu County. Inappropriate use of antimicrobials in poultry feed for growth promotion, product improvement, and non-adherence to withdrawal periods during antimicrobial treatment, challenge the ethics of antimicrobial use in poultry production. Low awareness and poor knowledge of antimicrobial residues among poultry farmers presents a significant issue contributing to antimicrobial resistance. This threatens public and environmental health due to the interconnectedness of humans and animals in the same ecosystem. The growing risks associated with antimicrobial resistance call for a One Health approach to mitigate AMR at the human, animal, and environmental interphase. In the framework of One Health, interventions are required to address the issue of AMR and inform policy directions towards ethical antimicrobial use and public health promotion. It is also of paramount importance that ethics ought to be a key factor of consideration in promoting good antimicrobial stewardship and fighting against AMR in poultry production.

## Data Availability

Data are available from the University of Nairobi repository URL:http://erepository.uonbi.ac.ke/

## Acknowledgements

The authors thank all the poultry farmers who participated in the study. A special thanks to the sub-county veterinary officers who granted access to the sub counties and made it possible for the study to be conducted. We express great gratitude to the livestock production officers for the immense support in mapping of the target poultry farmers, and Mr. Kelvin Ndeto, who assisted during data collection.

